# Machine learning-based predictive clinical model for *Shigella* spp. infection in children with diarrhea

**DOI:** 10.64898/2026.07.06.26357284

**Authors:** Francisco Sousa Junior, Jose Quirino Silva Filho, Alexandre Havt Binda, Gagandeep Kang, Margaret N. Kosek, Pascal O. Bessong, Amidou Samie, Rashidul Haque, Estomih R. Mduma, Jose P. Leite, Ladaporn Bodhidatta, Najeeha T. Iqbal, Nicola Page, Ireen Kiwelu, Zulfiqar A. Bhutta, Tahmeed Ahmed, Elizabeth Rogawski McQuade, James A. Platts-Mills, Eric R. Houpt, Aldo A. M. Lima

## Abstract

Diarrheal disease remains a significant cause of morbidity and mortality in children under five years of age in low-and middle-income countries. Identifying the etiology of diarrheal episodes represents a significant challenge in contexts with limited access to laboratory diagnostic methods, where therapeutic decisions are often based solely on clinical criteria, such as the presence of blood in the stool recommended by the World Health Organization (WHO). In this scenario, identifying combinations of clinical signs and symptoms could contribute to more precise therapeutic decisions. In this study, we developed and internally validated a predictive model based exclusively on clinical variables to identify episodes attributable to *Shigella* spp. infection, using as a reference an etiological outcome defined by quantitative molecular methods (qPCR). Secondary data from the Malnutrition-Enteric Diseases (MAL-ED) cohort, a multicenter study conducted in eight countries (2009–2016), with longitudinal follow-up of 1,715 children, were used. Diarrheal episodes were reconstructed from a disease surveillance database. Subsequently, fecal samples were temporally linked to these episodes, allowing the incorporation of molecular etiological data, defining diagnostic positivity for *Shigella* spp. After eligibility criteria and data processing, a final analytical database was obtained with 3,342 episodes and nine clinical variables (age, sex, blood in stool, bowel movement frequency, diarrhea duration, dehydration, fever, vomiting, and hospitalization) selected after multicollinearity assessment. Five machine learning algorithms were evaluated, with performance estimated by internal validation. Logistic regression showed the best discrimination (AUC = 0.789) and good calibration (Brier score = 0.077). At a cutoff point of 0.46, the model achieved a sensitivity of 0.753 and a specificity of 0.708. In comparison, the WHO score showed inferior performance (AUC = 0.556; sensitivity = 0.147; specificity = 0.965). The high negative predictive value (0.96) highlights the model’s ability to exclude cases not attributable to *Shigella* spp., suggesting potential utility as a tool to support primary clinical diagnostic decision-making and rational use of antibiotics in resource-limited settings.

## Introduction

Diarrheal disease remains among the leading causes of morbidity and mortality in children under five years of age in low-and middle-income countries [28]. Recent estimates from the Global Burden of Disease Study indicate that diarrhea remains among the leading causes of mortality in children under five years of age globally [8]. Although mortality has decreased in recent decades, morbidity remains high, being associated with impairments in linear growth, neurocognitive development, school performance, and reduced long-term economic productivity, creating cycles of poverty and social inequality in vulnerable communities [5,11], particularly when recurrent episodes occur in the first years of life [16]. Beyond the direct health consequences, diarrhea has profound social and economic impacts. Malnutrition caused by recurrent episodes impairs physical and cognitive growth, contributing to 15% of cases of stunting in children globally [5]. Furthermore, clinical practice in low-resource settings often faces difficulties in adequately identifying the causative agents of diarrhea [14]. And therapeutic decisions are frequently based on clinical markers such as the presence of blood in the stool, as recommended by both international and national guidelines for dysentery identification [29,23]. The unavailability of sensitive laboratory tests favors the empirical use of antimicrobials in contexts of diagnostic uncertainty. The lack of accurate diagnostic tools leads to overuse of antibiotics, contributing to the increase in antimicrobial resistance – a global public health problem. Studies [21,22] suggest that up to 92% of antibiotic treatments for childhood diarrhea are inappropriate, reinforcing the need for evidence-based strategies to guide the treatment of the disease. In this context, identifying the etiology of diarrheal episodes remains a challenge in settings with limited access to laboratory diagnostic methods. Among bacterial agents, *Shigella* spp. stands out as an important cause of inflammatory diarrhea and greater clinical severity in young children. Recent studies have reanalyzed data from the MAL-ED cohort study (Etiology, Risk Factors and Interactions of Enteric Infections and Malnutrition and the Consequences for Child Health and Development), conducted in eight countries (Dhaka, Bangladesh; Vellore, India; Bhaktapur, Nepal; Naushero Feroze, Pakistan; Venda, South Africa; Haydom, Tanzania; Fortaleza, Brazil; and Loreto, Peru) between 2009 and 2016 on childhood diarrhea in low-resource countries, using quantitative PCR to identify pathogens [21]. The study also highlighted the inaccuracy of using blood in stool as the sole indicator of *Shigella* spp., proposing a predictive score that includes other clinical characteristics to improve diagnosis and treatment. The study compared two methods for predicting and diagnosing shigellosis (infection caused by the bacteria *Shigella* spp.), analyzing two main criteria: sensitivity and specificity. The first approach used a predictive scoring system that showed a sensitivity of 50.4% and a specificity of 84.0%. This means that the method correctly identified about half of the actual cases of shigellosis (sensitivity), while avoiding false diagnoses in 84% of uninfected individuals (specificity). The second approach, based on current WHO clinical guidelines, recommends treating only subjects who present with bloody diarrhea. This strategy, although quite specific (96.5%), revealed a very low sensitivity of only 14.5%, correctly identifying a small fraction of shigellosis cases [21]. These results highlight the limitations of the traditional approach in the correct diagnosis of shigellosis, which is effective in avoiding false diagnoses but fails to recognize most actual infections, potentially leaving many individuals without adequate treatment. On the other hand, the scoring system proposed in the MAL-ED cohort study proved to be more balanced, significantly increasing the detection of shigellosis cases without drastically compromising the accuracy in excluding false positives. The application of a score-based predictive method proposed in the MAL-ED cohort [21] shows promise as an alternative. It improves diagnostic capability by increasing the identification of real cases, which can contribute to more assertive clinical decisions. However, despite its greater accuracy, molecular diagnosis (Gold standard) remains limited in low-resource settings due to costs, laboratory infrastructure, and processing time. This diagnostic gap creates a mismatch between widely available, but imprecise, clinical rules and more accurate, but less accessible, molecular methods. In this context, probabilistic approaches based on machine learning may represent a promising methodological alternative [27]. Supervised models allow the integration of multiple routinely collected clinical variables, capturing multivariate patterns associated with diarrhea attributable to *Shigella* spp., and approximating molecular classification from clinically available data [13]. In this study, we developed and internally validated a machine learning (ML)-based clinical probabilistic predictive model to identify diarrhea attributable to *Shigella* spp. in children under five years of age, using as a reference an outcome defined by a quantitative molecular pattern (AFe ≥ 0.5). The attributable fraction (AFe) represents an estimate of the etiological contribution of a specific pathogen to an episode of diarrhea, derived from statistical models based on the pathogen load detected by qPCR [15]. ML algorithms are capable of handling complex datasets, analyzing multiple clinical variables to identify patterns associated with different etiologies of diarrhea. To this end, we adopted an analytical strategy structured in three stages: (I) development of supervised models using nine clinical variables for the diagnosis of *Shigella* spp. (age, sex, presence of blood in stool, bowel movement frequency, duration of diarrhea, dehydration, fever, vomiting, and hospitalization) selected after multicollinearity assessment; (II) comparison of the performance of these models with the World Health Organization clinical rule based on a single marker; and (III) further development of models employing the same set of seven clinical variables for the diagnosis of *Shigella* spp. (presence of blood in stool, fever, prolonged duration, dehydration, vomiting, and severe frequency) previously used in analyses of the MAL-ED study, which allowed for the evaluation of the consistency and robustness of predictive performance in different variable specifications. This approach integrated epidemiological rigor in the definition of *Shigella* spp., from the molecular outcome to contemporary statistical modeling and machine learning techniques, with the aim of estimating the individual probability of shigellosis from routinely available clinical variables. By structuring explicit comparisons between probabilistic models and the traditional clinical rule, we sought to evaluate the potential for improvement in the diagnostic discrimination of *Shigella* spp., and its implications for the rational use of antimicrobials in resource-limited settings (Figure 5).

## Methods

### Study design and data source

This study followed an analytical observational design using data from the MAL-ED database, encompassing a wide range of information necessary for microbiological and clinical analysis, including 1715 children followed from birth to 24 months of age, with active home surveillance for episodes of diarrhea and systematic collection of fecal samples. The construction of the analytical database for diarrhea episodes was carried out by integrating two main data sources from the MAL-ED study: (1) the disease surveillance database (Illness) with 1,380,929 records and 19 variables and (2) fecal samples (dbtac) with 42,488 records and 18 variables, containing clinical and epidemiological information associated with the collections. Diarrhea episodes were defined using the standardized longitudinal structure in the Illness database, which allowed the explicit construction of the temporal windows of each episode (start and end), ensuring consistency with the standardized clinical definition of the MAL-ED study [21]. This approach was adopted to avoid pseudo-replication problems—that is, considering multiple days belonging to the same episode as independent observations—and to mitigate temporal dependence inherent in data organized at the daily level. The variable “depi3” (0 = outside of episode, 1 = within episode, 2 = episode start) was used to identify episodes, according to the MAL-ED definition of episodes separated by at least 48 consecutive hours without diarrhea, considering days with missing data as diarrhea-free. Episode identifiers were generated by cumulatively summing the onset events (depi3 = 2) for each individual, ensuring temporal consistency and alignment with the original study design [21]. The clinical variables considered included the presence of blood in the stool, fever, dehydration, episode duration, frequency of bowel movements, vomiting, and hospitalization. Episode-level variables, including duration, number of days with diarrhea, and clinical characteristics, were derived by aggregating the daily records. Thus, the analytical unit of the study was defined at the episode level, not the day level, ensuring independence between observations and statistical validity of subsequent analyses. For each sign or symptom (e.g., blood in stool, fever, vomiting, and dehydration), its occurrence on at least one day of the episode was considered, using a presence rule (maximum value observed during the period). Additionally, the number of days with available observation (saf_days_obs) was calculated, allowing for an assessment of the completeness of clinical information during the episode. This aggregation strategy can be interpreted as a temporal reduction process, in which longitudinal information is synthesized into variables representative of the clinical episode. This approach is widely used in clinical predictive models based on longitudinal data, as it preserves the clinical relevance of signs and symptoms without introducing dependence between observations [43]. This procedure constitutes a form of temporal reduction of longitudinal data, allowing the construction of independent clinical units for predictive analysis. The final result was a database with 8,758 episode records and 19 variables. After constructing the clinical episodes, the etiological data from fecal samples were linked, with 42,488 records for each episode. Initially, an expanded time window of ±7 days was applied in relation to the clinical interval of the episode, defined from the start and end dates of each episode. This approach was used to capture fecal samples potentially related to the clinical event, considering possible variations in the time of collection in relation to the course of the episode. Next, a filtering criterion was applied to the fecal samples based on the sample type (stool type), keeping only samples eligible for etiological analysis (e.g., M1 type samples, as defined by the MAL-ED study). Samples collected within the expanded window of distinct episodes were treated to avoid overlap or duplication of attribution, ensuring consistency in linking samples and episodes. After the filtering step, the final linking of fecal samples to episodes was performed considering only samples whose collection date fell within the clinical interval of the episode (between the start and end dates). This strategy ensures high specificity in etiological attribution, reducing the risk of incorrect association between exposure and outcome. The total number of fecal samples was 40,634 records; this process removed 1,854 samples. From this integration, variables from the fecal sample database were incorporated at the episode level, including the etiological outcome based on molecular methods, represented by the attributable fraction (AFe) for *Shigella* spp. The integration between the clinical episode database and the filtered fecal sample database resulted in an analytical database at the episode level containing 8,758 records and 37 variables. Linkage was performed based on the participant identifier (PID) and the temporal correspondence between the sample date and the clinical interval of the episode. From this integrated database, a database of all identified episodes used for descriptive and exploratory analyses was created; and a restricted database composed only of episodes that met the operational definition of diarrhea, according to established clinical criteria. This restricted database was used as the main set for the development and evaluation of predictive models.

### Etiological definition based on molecular quantification

Initially, typological consistency of the necessary quantitative variables was ensured by explicitly converting them to numerical format. A new binary variable, diarrhea, was then operationally defined, aligned with the clinical criteria of the MAL-ED study [21], as the presence of ≥3 liquid bowel movements in 24 hours (maxls ≥ 3) or the presence of visible blood in the stool (maxb ≥ 1). This definition allowed each record to be classified as diarrheal (1) or non-diarrheal (0), maintaining consistency with the previously described epidemiological structure [21]. Subsequently, the dependent variable shigella_pos was constructed, representing episodes attributable to Shigella/Enteroinvasive Escherichia coli (EIEC). This variable was defined as a value equal to 1 when two conditions were simultaneously met: (1) the record was classified as diarrheal and (2) the fraction attributable to the episode (AFe) for *Shigella* spp. was ≥ 0.5. Records that did not meet both criteria were classified as 0. Thus, the final outcome incorporates both the clinical definition of the event and the quantitative etiological inference derived from qPCR. The remaining records remained classified as not attributable to *Shigella* spp. Compared to the previous version of the database (37 variables), two new derived variables were added: diarrhea and shigella_pos. The inclusion of these two variables formalizes the definition of the supervised etiological outcome and concludes the epidemiological preparation stage of the database. This consolidation ensures that the database for final analysis is temporally consistent, coherent with the previously established etiological score of the MAL-ED cohort study. With the explicit definition of the binary outcome, the dataset proceeded to the modeling stage with the selection of clinical predictors and the development of molecular modeling (MM) models. This stage consolidates the previously described methodological sequence—identification of episodes, expansion of time windows, application of a filter to non-diarrheal samples, and preservation of molecular integrity in the diagnosis of *Shigella* spp.—culminating in the formal definition of the binary outcome that will be used in comparative analyses and MM models. Therefore, the final database comprised 8,758 episode records and 39 variables, of which 3,407 (38.9%) corresponded to samples on the initial day of a diarrheal episode (D1) and 5,351 (60.09%) to non-diarrheal samples (Figure 6). The molecular outcome identified 320 records attributable to *Shigella* spp. (3.65% of the total), corresponding to 9.39% among diarrheal episodes.

### Selection of Clinical Variables

After consolidating the final analytical database, it was decided that the modeling base would consist solely of cases of diarrhea episodes. A new database was then generated with a total of 3,407 records and 39 variables. This initiated the stage of carefully selecting candidate variables for the development of supervised models. The objective was to identify a set of clinical predictors capable of estimating the probability of *Shigella* spp. infection attributable to the diarrhea episode, while maintaining a strict separation between input variables and the etiological outcome [21]. Initially, laboratory information used exclusively to define the molecular outcome was excluded, ensuring that the model learned only clinical patterns associated with etiological inference, without incorporating variables derived from the reference pattern itself. This strategy was adopted to avoid information leakage and ensure inferential validity. Subsequently, an additional dimensional reduction stage oriented towards predictive modeling was performed. From the created database, only clinically interpretable variables potentially available at the time of the subject’s assessment in a care setting were retained. Redundant structural columns, administrative variables, intermediate fields from laboratory processing, and information unrelated to the supervised task were removed. After checking the consistency of the data, it was found that the variables *shigella_eiec_afe* and *shigella_eiec* had 65 missing values. It was decided to exclude them. As a result of this process, the dataset was reduced from 3,407 episodes to 3,342 episodes, fully preserving the number of observations and retaining only the predictors relevant to the modeling stage. In order to assess the structural independence between the candidate clinical predictors and reduce informational redundancy, a linear correlation analysis was conducted using Pearson’s coefficient (r). The correlation matrix was calculated considering the selected continuous and binary variables, assuming a linear approximation suitable for exploratory screening purposes. A threshold of |r| ≥ 0.70 was adopted as the operational criterion for multicollinearity. Variables that showed a correlation greater than this value with another predictor were evaluated for clinical and statistical redundancy. From this screening, the initial set of 39 variables was reduced to 9 clinically independent predictors (age, sex, presence of blood in stool, bowel movement frequency, diarrhea duration, dehydration, fever, vomiting, and hospitalization), all showing correlation coefficients below 0.30 among themselves. This procedure aimed to: (a) Minimize parametric instability in linear models; (b) Reduce the risk of overfitting due to informational redundancy; and (c) Preserve the clinical interpretability of the final model. The final selection included clinical variables directly observable in the care setting. These variables reflect the operational clinical phenotype of diarrhea and are aligned with the literature of the MAL-ED study [21]. The consolidated final analysis database comprised 3,342 records of diarrhea episodes and 9 variables, fully preserving cases eligible for etiological inference. This restriction is conceptually consistent with the study’s objective, since the etiological attribution to *Shigella* spp. It is defined only in the context of diarrhea episodes [21], and it is not epidemiologically appropriate to include non-diarrheal samples in the etiological modeling stage.

### Development and validation of predictive models

Five supervised algorithms with different structural assumptions were evaluated, aiming to compare linear and non-linear models and assess the robustness of predictive performance. The following Machine Learning classifiers were implemented: (1) Logistic Regression with L2 regularization [34]: a parametric linear model that assumes a linear relationship in the logit between predictors and outcome. L2 penalty was applied to reduce the risk of overfitting and stabilize coefficients in the presence of residual correlation between variables. This model was considered an interpretable reference (clinical baseline); (2) Random Forest [35]: an ensemble learning method, in which multiple base models (decision trees) are combined to produce a final prediction, a strategy that tends to reduce variance and improve predictive robustness; (3) Histogram-Based Gradient Boosting (HistGB) [36,37]: a boosting algorithm that builds sequential trees by optimizing gradients of the loss function, suitable for modeling complex relationships between clinical variables; (4) Support Vector Machine [38] with radial kernel (RBF): margin-maximization-based model, using nonlinear transformation to capture complex decision boundaries; (5) k-Nearest Neighbors (kNN) [39]: nonparametric method based on proximity in the feature space, without presupposing a specific functional form. The choice of these algorithms was based on their consolidated use in classification tasks in epidemiology and data science applied to health. The analytical unit was the individual diarrheal episode. The supervised outcome used in the modeling was the variable shigella_pos. Only clinical variables available at the time of the initial assessment were considered as predictors, ensuring applicability in clinical deployment scenarios. The variables were converted to an appropriate numerical format and organized into a predictor matrix (X) and a binary response vector (y). Data processing was performed using a reproducible analytical workflow using the scikit-learn library, a widely used set of tools in Python for developing and evaluating predictive models [40]. This procedure was performed within the same analytical workflow used to build the models, in order to prevent information from the assessment set from influencing the model training. To evaluate model performance and reduce the risk of overestimating results, an internal validation strategy was used based on repeatedly dividing the database into independent subsets. In each repetition, the models were developed using most of the data and evaluated on a separate subset that had not been used in training. This procedure allowed each episode to be evaluated by a model that had not had prior access to that observation, providing more robust estimates of predictive performance. Three analytical scenarios were evaluated. In the first, the predictive models were built using nine clinical variables selected after correlation analysis between variables. In the second, the models’ performance was compared with the clinical rule currently recommended by the World Health Organization, based on the presence of visible blood in the stool. In the third scenario, the models were re-evaluated using the same set of seven clinical variables previously used in the clinical score derived from the MAL-ED study, allowing for the evaluation of the consistency of the results under a comparable informational framework. The agreement between the risks estimated by the models and the observed frequencies of infection was assessed using calibration curves [33] and the Brier score [4]. The potential clinical utility of the model was explored through decision curve analysis, which estimates the net clinical benefit of the model at different probability levels used to guide therapeutic decisions. Additionally, a continuous reclassification measure was calculated to assess whether one model showed an improvement in risk classification compared to another.

## Statistical Analysis

The performance of the models in distinguishing between episodes attributable and not attributable to *Shigella* spp. infection was evaluated by the area under the ROC curve (AUC) (Figure 1), a widely used measure to quantify a model’s ability to differentiate individuals with and without the outcome of interest. The models were evaluated through internal validation based on independent subdivisions of the database, so that each episode was evaluated by a model trained on a different subset of data. 95% confidence intervals for the AUC were estimated using the DeLong method [6], and differences between ROC curves were compared using the DeLong test for correlated data. Sensitivity, specificity, positive predictive value (PPV) and negative predictive value (NPV), as well as balanced accuracy, were calculated at the cutoff point that provided the best balance between sensitivity and specificity. The observed prevalence of infection in the sample was considered in the calculation of predictive values. The agreement between the risks estimated by the model and the observed frequencies of infection was assessed using the Brier score [4] and calibration curves, according to recommendations for evaluating clinical prediction models [24,25]. Initially, it was observed that the probabilities estimated by logistic regression were not perfectly aligned with the observed risk. To improve this fit, a probability recalibration procedure was applied, using a flexible approach widely employed to adjust probabilistic predictions. This process was carried out within the same internal validation scheme used in the model performance evaluation [32]. The level of statistical significance was set at 5%, when applicable. All analyses were conducted in a Python environment (version 3.12.0), using the scikit-learn [40] and pandas [41] libraries.

**Figure 1.**
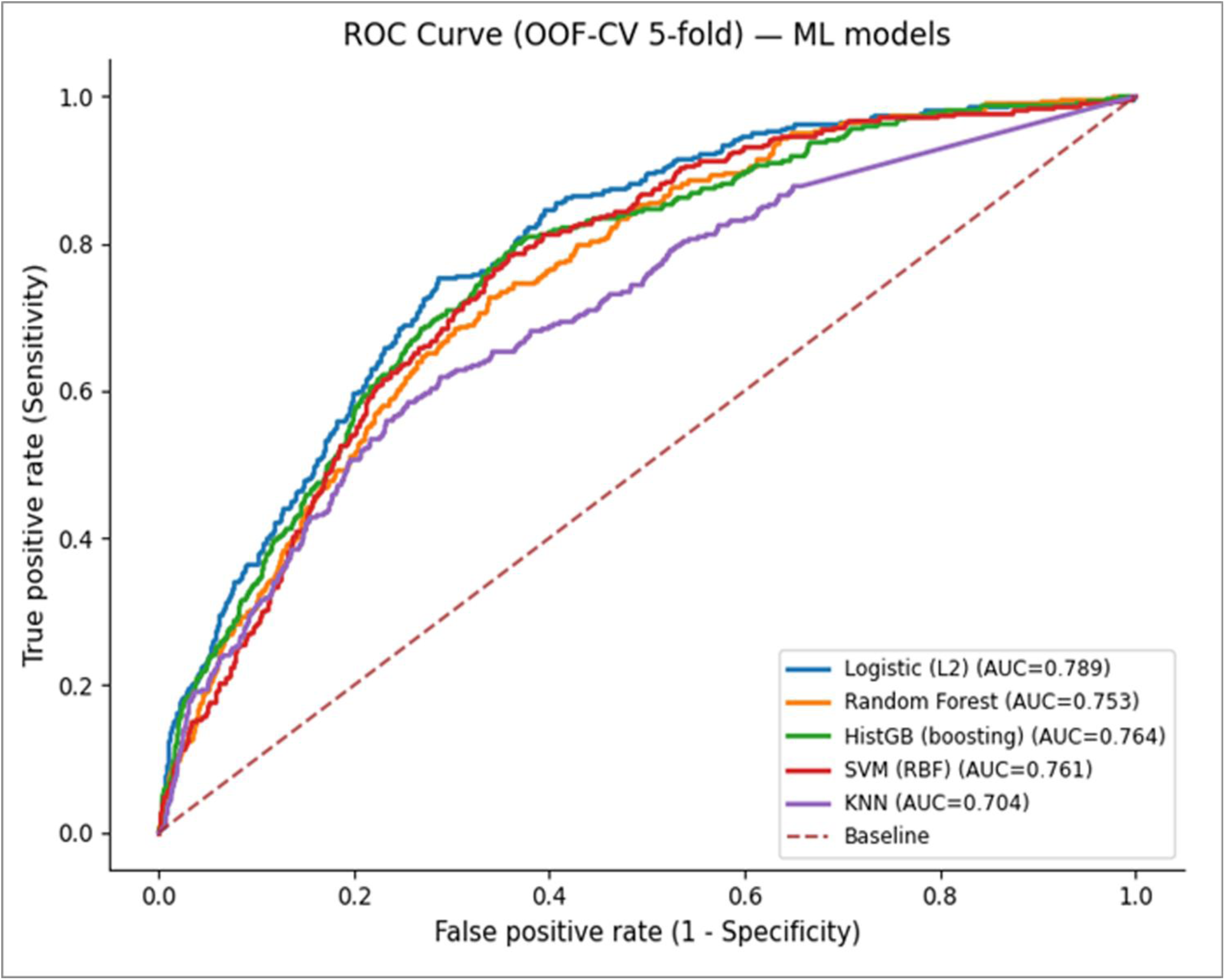
ROC curves of supervised models using nine clinical variables. ROC (Receiver Operating Characteristic) curves comparing the discriminatory performance of five supervised models for predicting diarrhea episodes attributable to *Shigella* spp. (episode attributable fraction ≥0.5). The models were developed using nine clinical variables available at the time of evaluation: age, sex, presence of blood in stool, bowel movement frequency, diarrhea duration, dehydration, fever, vomiting, and hospitalization. Performance was estimated by internal validation based on independent subdivisions of the dataset. Results are presented for logistic regression with L2 regularization, Random Forest, Gradient Boosting, Support Vector Machine, and k-Nearest Neighbors. The diagonal line indicates the expected performance for a random classifier (AUC = 0.5).

## Results

Among the clinical predictors evaluated (**Table 1**), the presence of blood in the stool showed the strongest association with episodes attributable to *Shigella* spp. (OR = 5.73; 95% CI 3.84–8.56). High bowel movement frequency (OR = 1.14; 95% CI: 1.07–1.21). Age and duration of diarrhea showed slight associations, reflecting the epidemiological pattern observed in multicenter studies of diarrheal etiology. Age and duration of diarrhea showed slight associations. Sex, fever, and dehydration did not show a significant association. On the other hand, episodes accompanied by vomiting (OR = 0.82; 95% CI: 0.60–1.12) and hospitalization (OR = 0.53; 95% CI: 0.06–4.56) showed a tendency toward an inverse association. The findings reinforce the clinical value of blood in the stool as the main indicator of *Shigella* spp. infection.

**Table 1.**
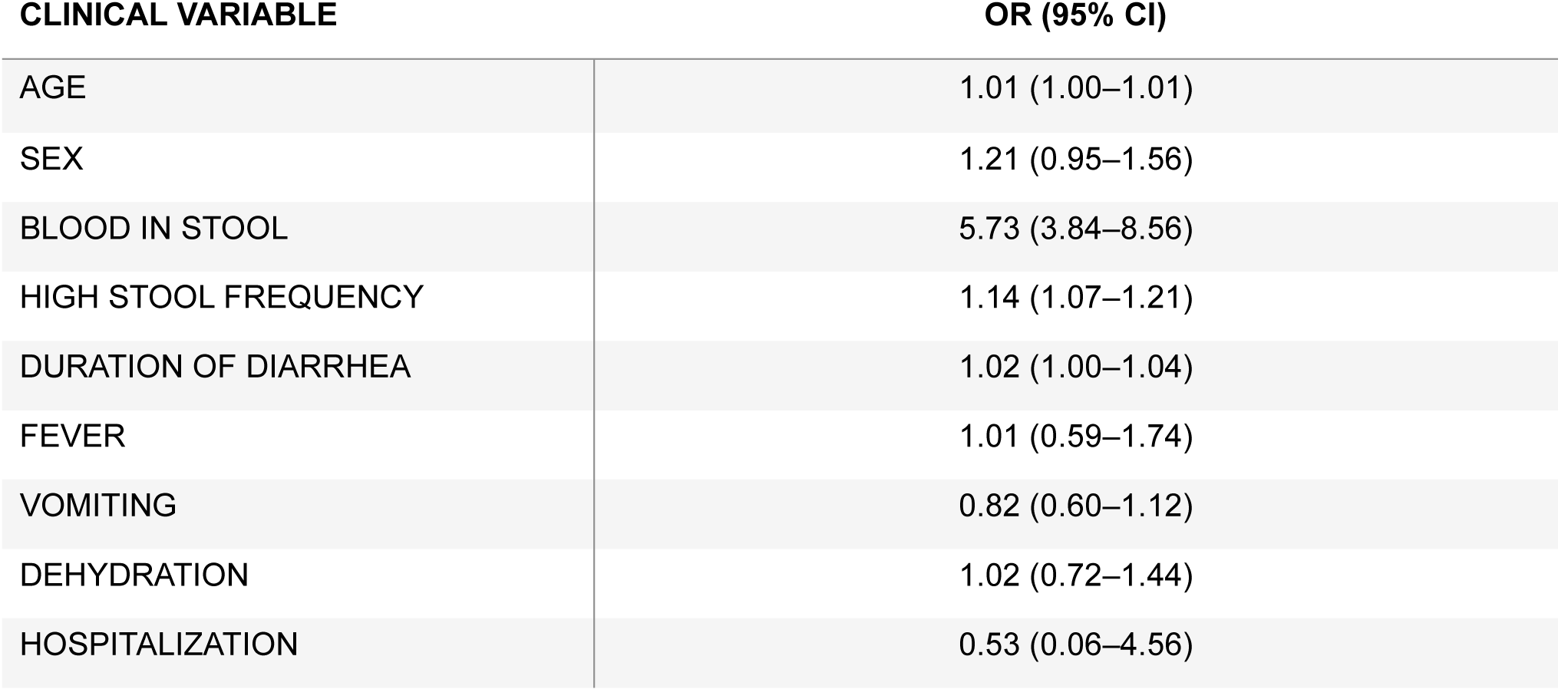
Clinical predictors associated with diarrhea attributable to *Shigella* spp. presents the clinical predictors associated with diarrhea attributable to *Shigella* spp. in a multivariable model. The presence of blood in the stool was the main predictor (OR = 5.73; 95% CI: 3.84–8.56), followed by high stool frequency (OR = 1.14; 95% CI: 1.07–1.21). Age and duration of diarrhea showed modest associations. Sex, fever, and dehydration did not show significant associations. Vomiting (OR = 0.82; 95% CI: 0.60–1.12) and hospitalization (OR = 0.53; 95% CI: 0.06–4.56) showed an inverse association trend. These findings reinforce the clinical value of blood in stool as the main indicator of infection by *Shigella* spp.

| CLINICAL VARIABLE | OR (95% CI) |
| --- | --- |
| AGE | 1.01 (1.00–1.01) |
| SEX | 1.21 (0.95–1.56) |
| BLOOD IN STOOL | 5.73 (3.84–8.56) |
| HIGH STOOL FREQUENCY | 1.14 (1.07–1.21) |
| DURATION OF DIARRHEA | 1.02 (1.00–1.04) |
| FEVER | 1.01 (0.59–1.74) |
| VOMITING | 0.82 (0.60–1.12) |
| DEHYDRATION | 1.02 (0.72–1.44) |
| HOSPITALIZATION | 0.53 (0.06–4.56) |

### Performance of Predictive Models

The prevalence of the outcome variable of *Shigella* spp. infection incidence was 9.39% (320/3,407). Model performance was evaluated by internal validation with data resampling, allowing different subsets of the sample to be used alternately for model training and evaluation. Among the classifiers evaluated, logistic regression showed the best discriminatory capacity (ROC-AUC 0.789; 95% CI 0.765–0.813), surpassing other machine learning models tested. All models showed discriminatory capacity superior to chance. Considering the low prevalence of the outcome, we also performed an additional analysis focused on the models’ ability to correctly identify positive episodes. Logistic regression showed the best performance in this analysis, with the highest F1-score (0.33), reflecting the balance between sensitivity and accuracy in identifying positive episodes, followed by other algorithms tested, including tree-based methods and support vector machines. **Table 2** summarizes the diagnostic performance of the evaluated models, including discriminatory capacity (AUC with a 95% confidence interval), sensitivity, specificity, and positive and negative predictive values. Performance was estimated by internal validation using different subsets of the database for model training and evaluation. Among the algorithms tested, logistic regression showed the best overall discriminatory performance, while some models showed higher sensitivity but lower specificity. Using the cutoff point that best balances sensitivity and specificity (0.50), logistic regression showed a sensitivity of 75.3% and a specificity of 71.4%, with a balanced accuracy of 73.4%. The positive predictive value was 21.8%, while the negative predictive value was 96.5%, indicating a high capacity of the model to correctly identify episodes not attributable to *Shigella* spp. infection. This high negative predictive value suggests potential clinical utility as a tool to support diagnostic exclusion in contexts with moderate disease prevalence. Initially, it was observed that the probabilities estimated by the model showed limited agreement with the observed frequencies. After calibration [31] adjustment performed using an isotonic regression method, there was a substantial improvement in this agreement, with a reduction in the overall probabilistic error measure (Brier score) from 0.188 to 0.077. The calibration curve demonstrated consistent alignment between estimated risk and observed risk after adjustment (**Figure 2**). In the clinical utility analysis, the calibrated model showed a positive net benefit across a wide range of probabilities used to guide therapeutic decisions, maintaining superior performance to the extreme strategies of treating all subjects or treating none, especially in low to intermediate risk scenarios (**Figure 3**). These results suggest the potential applicability of the model as a clinical risk stratification tool. Initially, a discrepancy was observed between the probabilities estimated by the model and the actual frequency of observed events.

**Figure 2.**
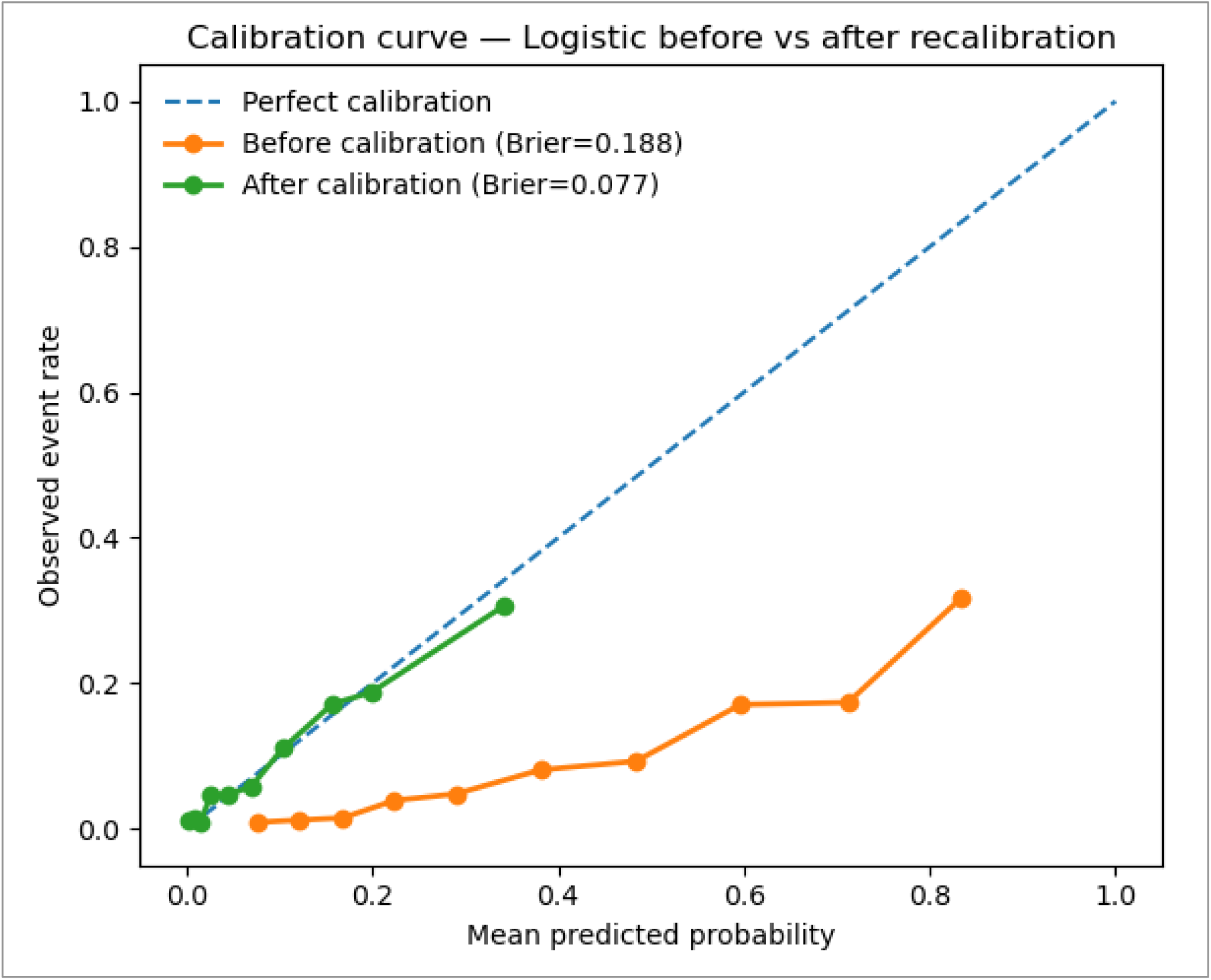
Calibration curve of logistic regression with nine clinical variables. Calibration curve evaluating the agreement between the probabilities predicted by the logistic regression model and the observed frequencies of episodes attributable to *Shigella* spp. The estimated probabilities were recalibrated using isotonic regression to improve the alignment between predicted risk and observed risk. The x-axis represents the probabilities predicted by the model and the y-axis represents the observed frequency of the outcome in each risk interval. The diagonal line represents perfect calibration. After recalibration, an improvement in the agreement between predicted and observed probabilities was observed, accompanied by a reduction in the Brier score from 0.188 to 0.077.

**Figure 3.**
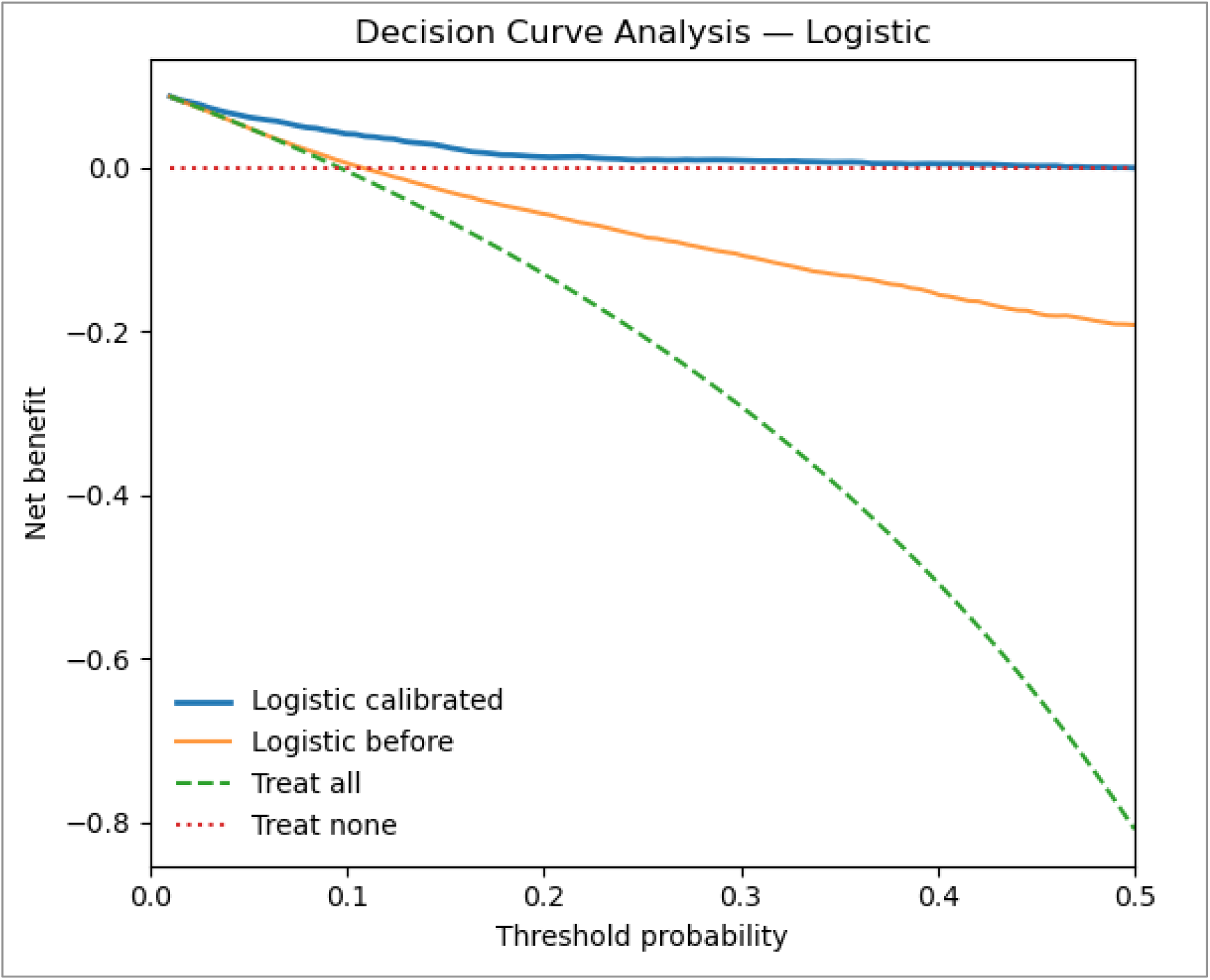
Decision curve analysis of the recalibrated logistic regression model. Decision curve analysis evaluating the potential clinical benefit of the recalibrated logistic regression model for predicting diarrhea episodes attributable to *Shigella* spp. The net benefit is presented on the y-axis as a function of the threshold probabilities used to guide therapeutic decisions on the x-axis. The model’s performance is compared with two reference strategies: treating all subjects and treating no subjects. The analysis demonstrates the model’s performance in different clinical risk scenarios considered relevant for therapeutic decisions in contexts with limited access to etiological diagnosis.

**Table 2.** Comparative performance of models for predicting diarrhea attributable to *Shigella* spp. (AFe ≥ 0.5). was constructed from predictive models developed in Python, using the Anaconda distribution (Anaconda Inc., Austin, TX, USA), in a Jupyter Notebook environment, with libraries such as pandas [41], NumPy [42], and scikit-learn [40]. Initially, clinical variables were transformed into binary and categorical predictors through feature engineering, a process of transforming raw variables into structured representations with the aim of improving model performance. Model performance was evaluated using stratified cross-validation (5-fold), in which the data were partitioned into subsets while preserving class proportions. In each iteration, models were trained on part of the data and evaluated on independent subsets not used for training. The area under the ROC curve (AUC) and its 95% confidence intervals were estimated using the DeLong method. For each model, the decision threshold was defined using the Youden index, from which sensitivity, specificity, positive predictive value, negative predictive value, and balanced accuracy were calculated.

| MODEL | AUC (95% CI) | SENSITIVITY | SPECIFICITY | PPV | NPV | BALANCED ACCURACY |
| --- | --- | --- | --- | --- | --- | --- |
| WHO CLINICAL RULE (BLOOD IN STOOL) | 0.556 (0.543–0.569) | 0.147 | 0.965 | 0.218 | 0.941 | 0.556 |
| MAL-ED CLINICAL SCORE (QPCR) | 0.783 | 0.500 | 0.840 | — | — | 0.670 |
| LOGISTIC REGRESSION (L2) | **0.789 (0.765–0.813)** | **0.753** | **0.714** | **0.218** | **0.964** | **0.733** |
| RANDOM FOREST | 0.753 (0.728–0.779) | 0.728 | 0.662 | 0.185 | 0.958 | 0.695 |
| GRADIENT BOOSTING | 0.764 (0.738–0.790) | 0.800 | 0.635 | 0.188 | 0.967 | 0.717 |
| SUPPORT VECTOR MACHINE | 0.761 (0.736–0.786) | 0.784 | 0.642 | 0.188 | 0.965 | 0.713 |
| K-NEAREST NEIGHBORS | 0.704 (0.673–0.734) | 0.618 | 0.711 | 0.185 | 0.946 | 0.665 |

**Abbreviations:** AUC = area under the ROC curve; 95% CI = 95% confidence interval; PPV = positive predictive value; NPV = negative predictive value.

### Clinical Predictors and Age-Stratified Sensitivity Analysis

In the multivariate analysis, the presence of blood in the stool (adjusted OR 5.73; 95% CI 3.84–8.56) and vomiting (adjusted OR 0.82; 95% CI 0.60–1.12) stood out as independent clinical predictors, while hospitalization did not show a statistically significant association. The permutation importance analysis identified age as the main determinant of overall discriminatory capacity. In the sensitivity test, the exclusion of the age variable resulted in a substantial reduction in the ROC-AUC (0.789 vs 0.558), evidencing a strong age contribution. In the age-stratified analysis, it was observed that, in infants (<12 months), the exclusion of age significantly reduced discrimination (AUC 0.763 versus 0.601). However, among children aged 12–23 months, performance also declines after age is removed (AUC 0.661 versus 0.601), indicating an independent contribution of clinical signs and symptoms in this population. In the 24–59 month group, the reduced number of cases limited the stability of the estimates.

### Comparison with the WHO clinical rule

The clinical rule recommended by the WHO, based exclusively on the presence of visible blood in the stool, showed substantially lower performance when compared to the supervised models. An AUC of 0.556 (95% CI 0.543–0.569) was observed, with a sensitivity of 0.147 and a specificity of 0.965. In contrast, all machine learning models using nine clinical variables showed AUCs greater than 0.70 (ranging from 0.704 to 0.789), with substantially higher sensitivity (≈0.61–0.80) and moderate specificity (≈0.64–0.71). Regularized logistic regression showed the best overall performance (AUC = 0.789; 95% CI 0.765–0.813). The difference between the AUCs of the logistic regression and the WHO rule was statistically significant according to the DeLong test (p < 0.001). The ROC curves show a clear separation between the multivariate approach and the univariate rule based exclusively on dysentery. At the cutoff point (0.50) that best balances sensitivity and specificity, the logistic model showed a sensitivity of 75.3% and a specificity of 71.4%, while the WHO rule showed a markedly lower sensitivity of 14.7%. Considering the prevalence of approximately 9.39%, the logistic model presented a negative predictive value of 0.964. This high negative predictive value indicates that, when the model classifies an episode as not attributable to *Shigella* spp., the residual probability of error is less than 4%. In contexts with limited access to molecular diagnostics, this exclusion capability has direct clinical relevance, as it allows for more reliable identification of episodes with a low probability of invasive bacterial etiology, potentially reducing unnecessary empirical antibiotic prescriptions and contributing to a more rational use of antimicrobials.

### Analysis using clinical variables from the MAL-ED score

As an additional analysis to allow methodological comparability with the clinical score developed in the MAL-ED study, the models were re-evaluated using the seven clinical variables (presence of blood in stool, fever, prolonged duration of diarrhea, dehydration, vomiting, high bowel movement frequency, and severity) previously adopted in the MAL-ED score. The results showed a substantial reduction in discriminatory performance compared to the scenario with nine variables. The ROC curves (**Figure 4**) demonstrated overall performance close to the non-discrimination line, with AUC values ranging approximately between 0.552 and 0.588, indicating limited ability to distinguish between episodes attributable and not attributable to *Shigella* spp. infection. Among the models evaluated, the Histogram-Based Gradient Boosting (HistGB) showed the best relative performance, although still modest (AUC≈0.588), and the other models showed similar performance. This convergence suggests that, in this scenario, the limitation is not in the algorithm, but in the low informational capacity of the set of predictors. Comparatively, the performance observed by the predictive model approaches that reported for approaches based exclusively on simplified clinical criteria, such as the World Health Organization (WHO) recommendation based on the presence of blood in the stool. This finding reinforces that the set of variables in the MAL-ED score, while clinically relevant, is insufficient to adequately capture the etiological complexity of diarrhea attributable to *Shigella* spp. when used in isolation in predictive models. The reduced performance compared to the expanded scenario suggests a critical role for additional variables, particularly age and sex, which can act as risk modifiers and contribute to clinical stratification. Taken together, these results indicate that models based exclusively on MAL-ED score variables have limited utility for individual etiological prediction and highlight the need for more comprehensive multivariate approaches.

**Figure 4.**
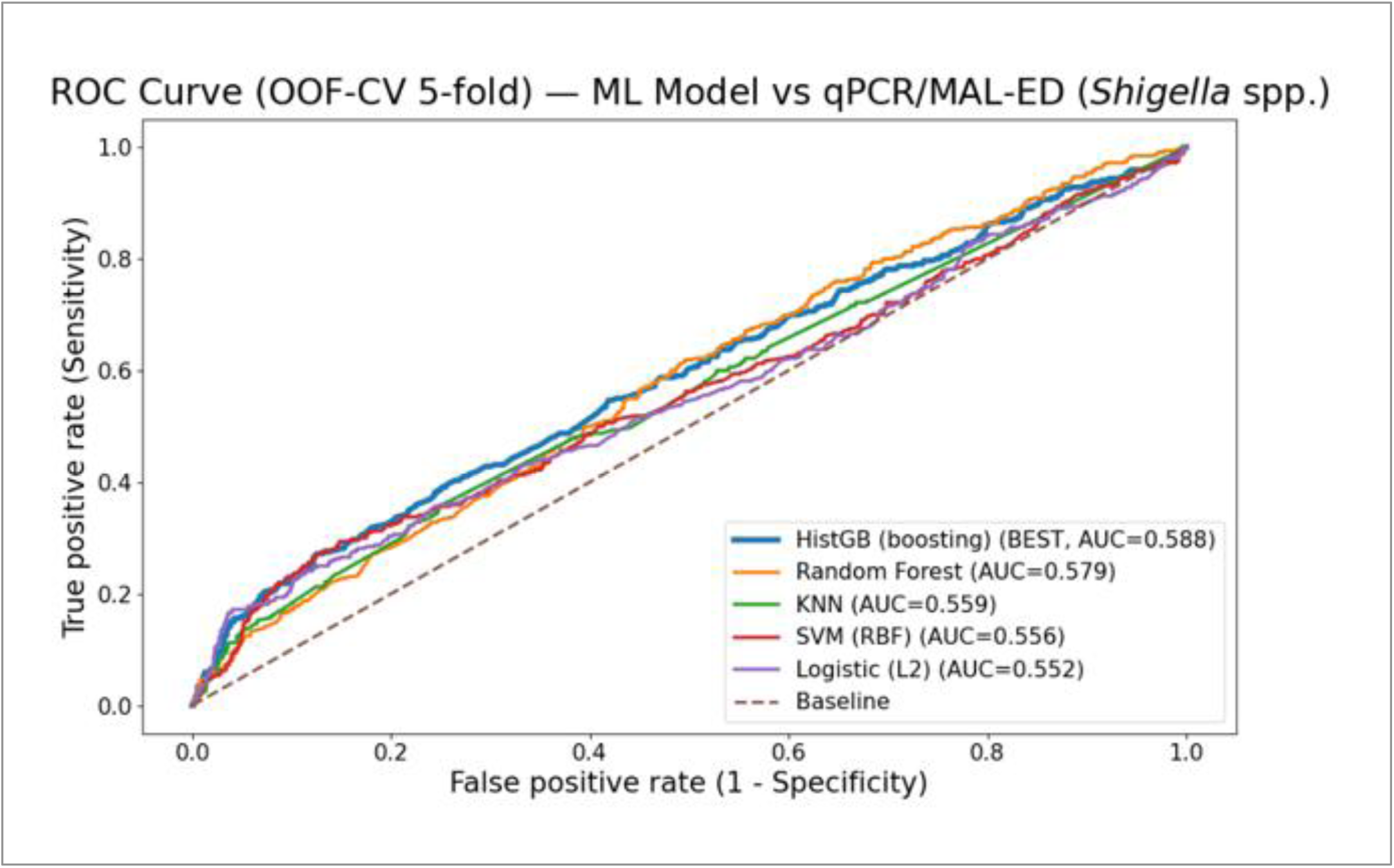
Performance of supervised models using the restricted set of clinical variables from the MAL-ED score. ROC curves of models trained with seven clinical variables (blood in stool, fever, diarrhea duration, dehydration, vomiting, bowel movement frequency, and severity), evaluated by internal validation. Logistic regression, Random Forest, Gradient Boosting, SVM, and k-NN showed similar and limited performance, with AUCs between ∼0.552 and 0.588 and curves close to the non-discrimination line. The findings indicate low predictive capacity for identifying episodes attributable to *Shigella* spp., lower than the scenario with a greater number of predictors and comparable to simplified clinical approaches.

**Figure 5.**
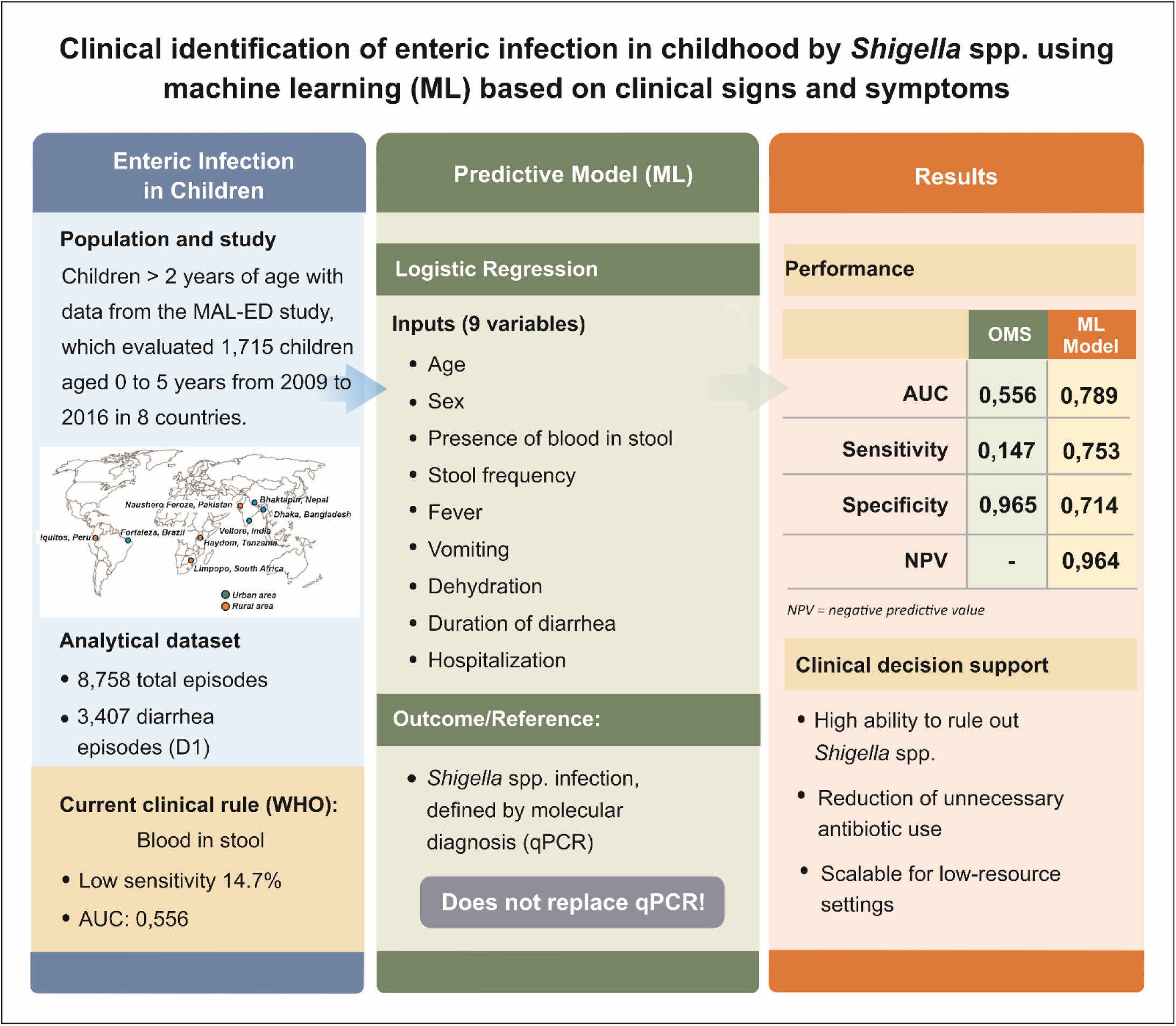
Clinical identification of enteric infection in childhood by *Shigella* spp. using machine learning from clinical signs and symptoms. The figure presents the methodological flow and the main findings of the study. The clinical rule currently recommended by the World Health Organization (WHO), based on the presence of blood in the stool, demonstrated limited performance, with low sensitivity (14.7%) and an area under the curve (AUC) of 0.556. Data from the MAL-ED study, involving 1,715 children aged 0 to 5 years followed between 2009 and 2016 in eight countries (Brazil, Peru, South Africa, Tanzania, India, Nepal, Pakistan, and Bangladesh), were analyzed. The analytical base comprised 8,758 total episodes, of which 3,342 corresponded to episodes of diarrhea on the initial day (D1), used as the main set for modeling. The predictive model was developed using logistic regression, utilizing nine clinical variables: age, sex, presence of blood in stool, bowel movement frequency, fever, vomiting, dehydration, diarrhea duration, and hospitalization. The etiological outcome was defined by molecular diagnosis (qPCR). The model showed superior performance to the WHO approach, with an AUC of 0.789, sensitivity of 0.753, specificity of 0.714, and a high negative predictive value (NPV = 0.964), indicating a strong ability to rule out *Shigella* spp. infection. Although it does not replace molecular diagnosis, the model demonstrates usefulness as a clinical decision support tool, contributing to the reduction of unnecessary antibiotic use and being potentially scalable to contexts with limited laboratory resources.

**Figure 6.**
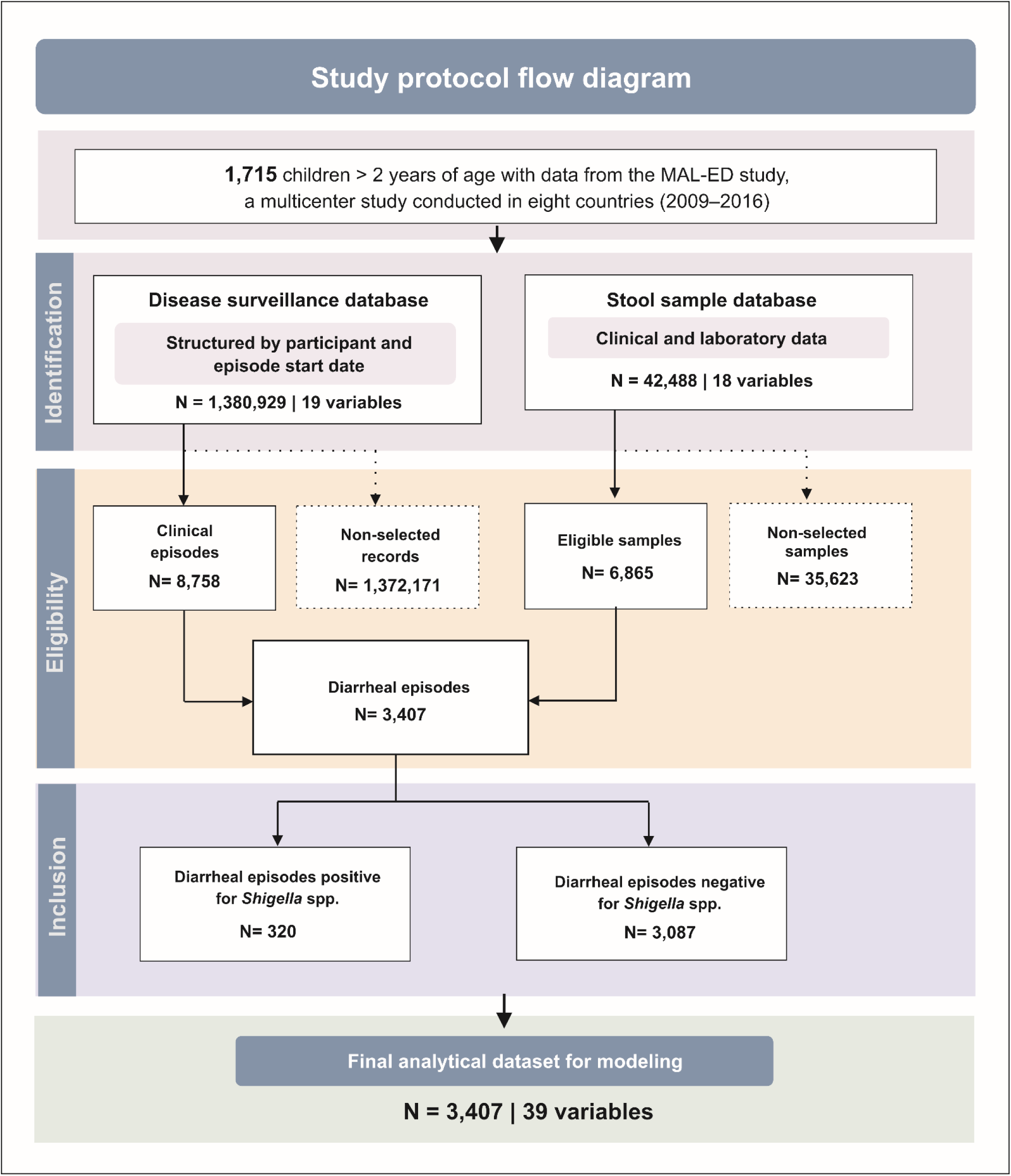
Flowchart of the study protocol and variable selection. The study integrated data from the MAL-ED cohort from the disease surveillance database (N = 1,380,929) and the fecal sample database (N = 42,488). 8,758 clinical episodes and 6,865 eligible samples were identified, which, after integration of the databases, resulted in 3,407 unique diarrheal episodes with linked data. The episodes were classified according to the presence of *Shigella* spp. based on quantitative molecular diagnosis (qPCR; AFe ≥ 0.5), totaling 320 positive and 3,087 negative episodes. The final analytical database included 3,407 episodes and 39 variables, forming the basis for the development of the supervised models.

## Discussion

The results of this study demonstrate that a multivariate probabilistic model trained with an outcome defined by a quantitative molecular pattern substantially outperforms the clinical rule currently recommended by the WHO for identifying episodes attributable to *Shigella* spp. The significant difference in discriminatory performance (AUC 0.789 versus 0.556), associated with a marked increase in sensitivity (0.753 versus 0.147) and a high negative predictive value (0.964), suggests that the structured integration of multiple clinical variables allows for a more precise capture of the clinical profile associated with invasive bacterial etiology. Recalibration of predicted probabilities substantially improved the probabilistic performance of the model, reinforcing the importance of the estimated absolute probability—and not just the relative ranking of individuals—in clinical decision-making. Decision curve analysis demonstrated incremental net benefit mainly at low to intermediate thresholds, consistent with outpatient settings where therapeutic decisions are made under diagnostic uncertainty. The decision curve suggests that recalibration enhances the model’s clinical utility by aligning estimated probabilities with observed risk, directly impacting the net benefit calculation. The greatest gain occurs at low to moderate thresholds—a typical range for empirical decisions in outpatient or resource-limited settings, where the acceptable risk for initiating antibiotics tends to be relatively low. Under these conditions, the calibrated model reduces unnecessary interventions compared to the treat-all strategy, while avoiding the undertreatment associated with the treat-none strategy. These findings indicate that proper calibration represents not only statistical refinement but also a determining component of the model’s clinical applicability. These results support the model’s role as a tool for risk exclusion and stratification, and not as a substitute for quantitative molecular confirmation. In resource-limited settings, where therapeutic decisions are often made in the absence of laboratory confirmation [30], the observed high exclusion capacity assumes direct clinical relevance. The high negative predictive value indicates potential usefulness as a decision support tool, reducing the likelihood of unnecessary empirical prescribing and potentially contributing to mitigating the selective pressure associated with bacterial resistance [30]. The clinical strategy recommended by the WHO, based on the presence of visible blood in the stool, has historically been driven by operational pragmatism [30]. However, evidence derived from quantitative molecular methods demonstrates that a substantial proportion of attributable episodes occur in the absence of classic dysentery [15,21]. In line with these findings, we observed a sensitivity of less than 20% for the WHO criterion, characterizing an essentially confirmatory and insensitive model for etiological screening. Sensitivity analysis revealed a strong contribution of age to overall discrimination, reflecting the epidemiological pattern of infection in the studied population. However, in the 12–23 month age group, clinical signs and symptoms maintained a relevant independent contribution, even after excluding the age variable, suggesting that the model integrates additional clinical information beyond the isolated age effect. The clinical score developed within the MAL-ED framework represented an important methodological advance [19,21]. The similar performance observed in this analysis suggests that the predictive gain stems predominantly from structured probabilistic modeling associated with rigorous internal validation, and not only from the expansion of the information set. The convergence of performance between regularized logistic regression and tree-based methods reinforces that the clinical approach associated with attributable shigellosis is robust and capturable by supervised modeling [7,9,12,17,26]. Some limitations should be considered. The model was only internally validated, and external validation is necessary to confirm generalization. The observed age contribution requires contextualized interpretation, and the reduced number of cases in older age groups limits specific inferences in these groups. Furthermore, the outcome based on AFE ≥ 0.5 assumes that quantitative attribution reflects individual clinical causality [15,21]. Despite these limitations, the findings suggest that incorporating probabilistic modeling based on molecular outcome may represent a viable intermediate step between traditional syndromic diagnosis and quantitative laboratory confirmation, particularly in contexts with limited access to qPCR.

## Conclusions and Future Extensions

This study demonstrated that etiological inference based solely on the presence of visible blood in stool, as recommended by the WHO, has limited discriminatory capacity to identify episodes attributable to *Shigella* spp. [20], when compared to molecular diagnosis. In contrast, multivariate approaches—including the MAL-ED clinical score and supervised machine learning models—show superior and consistent performance. This predictive gain reflects the ability to integrate multiple clinical signs and symptoms into individual risk estimates. While not replacing molecular diagnosis, the proposed approach translates the etiological definition into a probabilistic estimate based on clinical data available at the point of care, representing a potentially scalable strategy to reduce diagnostic gaps in resource-limited settings and promote the rational use of antibiotics, preventing the escalation of pressure on resistance to these drugs.

## Author Contributions

AAML, JQSF, and FSJ contributed to the study conception, methodological design, and data collection. FSJ performed data curation, statistical analysis, implementation of machine learning models, and drafted the initial version of the manuscript. AAML and JQSF contributed to the theoretical framework and critically revised the manuscript for important intellectual content. AH, GK, MNK, POB, AS, RH, ERM, JPL, LB, NTI, NP, IK, ZAB, TA, ETRM, JAPM, and ERH contributed to data acquisition, project supervision, and provided critical revisions of the manuscript for important intellectual content. All authors contributed to data interpretation, reviewed and approved the final version of the manuscript, and agree to be accountable for all aspects of the work. AAML is the corresponding author.

## Availability of data and materials

The data supporting the conclusions of this study are available upon reasonable request to the corresponding author. The machine learning model described in this work is in the process of intellectual property registration, which may imply partial restrictions on access to the source code and certain methodological components.

## Ethics approval and consent to participate

The MAL-ED study protocol and informed consent form were approved by the Brazilian National Commission for Ethics in Research (CONEP), under the National Health Council of the Ministry of Health, under registration number 15701 (Opinion No. 232/10; Process No. 25000.639836/2009-76), and by the Research Ethics Committee of the Federal University of Ceará (COMEPE/UFC) under registration No. 246/09. The study was conducted in compliance with Brazilian regulations for research involving human subjects, including National Health Council Resolution No. 196/96 and complementary guidelines. Further details of the MAL-ED study have been previously described (Murray-Kolb et al., 2014).

## Consent for publication

Consent for publication: not applicable.

## Financing

The Article Processing Charge (APC) for this publication was funded by the Coordination for the Improvement of Higher Education Personnel (CAPES). This work was carried out with the support of the Coordination for the Improvement of Higher Education Personnel - Brazil (CAPES) - Funding Code 88887.175863/2025-00.

## Acknowledgments

For open access purposes, the authors have applied a Creative Commons Attribution (CC BY) license to any version of the manuscript accepted for publication (Author Accepted Manuscript) resulting from this submission. During the preparation of this manuscript, the authors used ChatGPT Plus (version 5.2, OpenAI) and Paperpal Prime (Editage, Cactus Communication Service Pte Ltd, Singapore) as editorial assistance tools to support the improvement of language, structural organization, text clarity, and formatting of manuscript sections. The artificial intelligence tool was not used to generate original scientific data, perform analyses, interpret results, or produce figures or tables. All experimental design, data collection, statistical analyses, interpretation of results, and scientific conclusions were developed and verified exclusively by the authors. The authors critically reviewed and edited all AI-assisted content and assume full responsibility for the accuracy, integrity, and originality of the manuscript.

## List of Abbreviations

5-Fold: Five-fold cross-validation
ML: Machine Learning
AUC: Area Under the Curve
Brier score: Measure of accuracy of probabilistic predictions (mean squared error of predicted probabilities)
CONEP: National Research Ethics Commission
COMEPE: Research Ethics Committee
dbtac: Database of TaqMan Array Card (TAC) molecular assays
DeLong: Statistical method for comparing correlated AUCs and estimating confidence intervals
Illness: Illness surveillance dataset
MAL-ED: Etiology, Risk Factors and Interactions of Enteric Infections and Malnutrition and the Consequences for Child Health and Development
maxb: Maximum number of days with visible blood in stool during the episode
ML Models: Machine Learning Models
NPV: Negative Predictive Value
OMS (WHO): World Health Organization
OOF-CV: Out-of-Fold Cross-Validation predictions
pid: Participant ID
PPV: Positive Predictive Value
qPCR: Quantitative Polymerase Chain Reaction
ROC curve: Receiver Operating Characteristic curve
saf_days_obs: Number of days with available surveillance (SAF) data within the episode
scikit-learn: Python machine learning library for predictive modeling
shigella_eiec: Detection of *Shigella*/EIEC by molecular assays
shigella_eiec_afe: Attributable fraction estimate for *Shigella*/EIEC per episode
TCLE: Informed Consent Form (Termo de Consentimento Livre e Esclarecido)

